# Strategies for Improving the Performance of Prediction Models for Response to Immune Checkpoint Blockade Therapy in Cancer

**DOI:** 10.1101/2023.07.07.23292316

**Authors:** Tiantian Zeng, Jason Zhang, Arnold Stromberg, Jin Chen, Chi Wang

## Abstract

Immune checkpoint blockade (ICB) therapy holds promise for bringing long-lasting clinical gains for the treatment of cancer. However, studies show that only a fraction of patients respond to the treatment. In this regard, it is valuable to develop gene expression signatures based on RNA sequencing (RNAseq) data and machine learning methods to predict patients’ response to the ICB therapy, which contributes to more personalized treatment strategy and better management of cancer patients. However, due to the limited sample size of ICB trials with RNAseq data available and the vast number of candidate gene expression features, it is challenging to develop well-performed gene expression signatures. In this study, we used several published melanoma datasets and investigated approaches that can improve the construction of gene expression-based prediction models. We found that merging datasets from multiple studies and incorporating prior biological knowledge yielded prediction models with higher predictive accuracies. Our finding suggests that these two strategies are of high value to identify ICB response biomarkers in future studies.

## Introduction

Immunotherapy has emerged recently as a promising and viable treatment option for many cancer patients [1]. Among multiple types of immunotherapy, the immune checkpoint blockade (ICB) therapy, which aims at blocking the interaction of inhibitory receptors expressed on the surface of immune cells [2], has been proved applicable in helping the immune system target and attack cancer cells [3, 4]. Particularly, ICB can provide exceptional clinical gains in the treatment of a handful cancer, melanoma, mostly because the spontaneous regression of melanoma is closely related to the immune response [5, 6]. Despite the success of ICB therapy in the treatment of melanoma, however, recent studies showed that only around one-third of patients would respond to the ICB therapy [7]. The reason for the heterogeneous response still remains unclear and requires further investigations [1, 8, 9].

It is therefore desired to develop biomarkers that can predict patient’s response to ICB therapy, which will contribute to better stratification of patients to maximize therapeutic benefit. Previous studies showed that tumor mutational burden, microsatellite instability are predictive biomarkers [10–12]. Gene expression signatures have also been demonstrated as valuable for predicting ICB treatment response in melanoma patients [13–17]. However, the sample sizes of ICB clinical studies that have gene expression profiling data available are very limited [13]. Table 1 listed three published studies, each of which had less than 60 patients. The lack of large scale datasets makes it challenging to construct reliable prediction models. Although an alternative approach of using data from patients without ICB treatment to develop immune response signatures and transfering the results to predict ICB treatment response has been proposed [13, 14], it is still highly desired if the signatures could be directly built on patients with ICB treatment. Further, gene expression profiling technologies, such as RNA-sequencing (RNA-seq), are powerful to simultaneously quantify more than 10,000 genes’ expression levels. It is challenging to identify informative gene features and their complex relationships to build accurate prediction models, especially when the sample size is small.

**Table 1.**
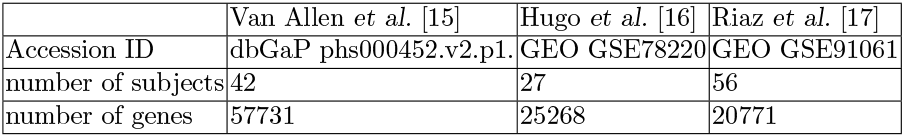
Datasets used in the study.

|  | Van Allen *et al.* [15] | Hugo *et al.* [16] | Riaz *et al.* [17] |
| --- | --- | --- | --- |
| Accession ID | dbGaP phs000452.v2.p1. | GEO GSE78220 | GEO GSE91061 |
| number of subjects | 42 | 27 | 56 |
| number of genes | 57731 | 25268 | 20771 |

In this paper, we investigated the potential of the following two strategies to enhance the development of gene expression signatures for ICB treatment effect prediction in melanoma patients. The first strategy is to merge data from different ICB clinical studies. Merging datasets has been shown as a viable approach to increase the sample size and thus improve the power of biomarker development in various biomedical applications [18, 19]. We explored the potential benefit of merging three published datasets [15–17] for the prediction of ICB treatment response. The second strategy is to leverage prior biological knowledge to use more informative and biologically relevant features for the construction of prediction models. It has been suggested that expressions of immune checkpoint genes and their interactions are relavent to tumor response to ICB therapy [13, 14]. We explored whether focusing on pairwise relation features among these immune checkpoint genes, as suggested by [14], could improve feature selection and prediction performance of the models.

## Materials and Methods

### Study design

Fig 1 presents an overview of our study design. Firstly, using each individual RNA-seq dataset, we leveraged prior biological knowledge and focused on immune checkpoint genes, where the pairwise relation of those genes were considered as candidate features for subsequent prediction model development. Secondly, we merged individual datasets to increase the sample size, where the presence of batch effect was assessed. Thirdly, based on the merged data, we built prediction models based on three commonly used machine learning algorithms, i.e. Random forest [20], Least absolute shrinkage and selection operation (LASSO) [21], and XGBoost [22]. Fourthly, we evaluated the performance of prediction models based on the receiver operating characteristic (ROC) curve and area under the curve (AUC) using cross-validation. To investigate the power of merging multiple datasets, we compared AUCs from models based on the merged dataset versus those based on each individual dataset. Besides, in order to investigate the benefit of incorporating prior information in feature selection, we compared AUCs from models built based on features characterizing pairwise relation of immune checkpoint genes versus those based on the original expression features of all genes.

**Fig 1.**
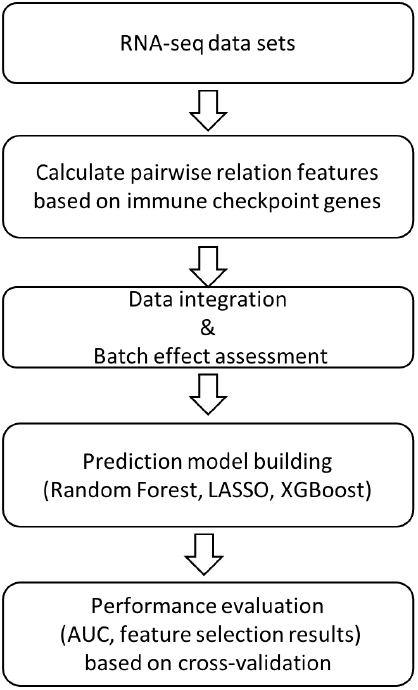
The flowchart of this study.

### Datasets

We considered three published melanoma datasets [15–17] as listed in Table 1. RNA sequencing data were downloaded in the unit of fragments per kilobase of transcript per million mapped reads (FPKM) or read counts, and transformed into the unit of transcripts per million (TPM). There were 18878 common gene features across the three datasets. The total sample size was 125. The detailed information of the three datasets is summarized in Table 1.

The response variable was defined to be a binary variable, ‘response’ or ‘non-response’ to the treatment. Since the response annotations for each dataset were not exactly the same, a standard classification used in this study was given following the definition from Auslander paper [14], where ‘complete response’, and ‘partial response’ were classified as ‘response’, and ‘nonresponse’, ‘progressive disease’, etc. were classified as ‘non-response’.

### Features Characterizing Pairwise Relations of Immune Checkpoint Genes

Due to the large number of gene features in one dataset, we focused on immune checkpoint genes and considered a similar set of candidate features as in [14]. Specifically, we considered 26 immune checkpoint genes that had been reported in the literature and were available in the RNA-seq datasets. As those immune checkpoint genes co-stimulatory and co-inhibitory, we considered pairwise relations between those gene expressions to capture their interactions [14, 23–26]. For a gene pair *x* and *y*, we define the following expression function was used:

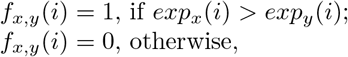

where *exp*_*x*_(*i*) and *exp*_*y*_(*i*) denote expressions of *x* and *y* in sample *i*. We further focused on the gene pairs containing at least one of the six genes, i.e. PD-1, PD-L1, CTLA-4, CD28, CD80 and CD86, that are directly associated with anti-CTLA-4 and anti PD-1 blockade therapy [27–29]. Hence, we obtained a total of 135 pairs forming candidate features for building prediction models.

### Data Integration

To integrate datasets from different sources, non-biological experimental variations, i.e. batch effects, should be evaluated [30]. Heatmaps and hierarchical clustering were used to visualize the batch effect, where samples from the same dataset clustered together would indicate the existence of a batch effect [31, 32].

### Prediction Model Building

We considered the following three frequently used statistical/machine learning methods to build models for predicting response to immune checkpoint blockade therapy based on features of the pairwise relations between immune checkpoint genes.

#### Random Forest

Random forest is a well established ensemble learning algorithm that can be applied for classification. It is formed by a large amount of individual decision trees, and then operates as an ensemble. Random forest applies a widespread technique of bagging, or called bootstrap aggregating while training the algorithm, but it includes implementing an essential modification of bagging in order to obtain an ensemble of de-correlated trees. [20] Package ‘randomForest’ in R (Version: 4.6-14) was used in this study.

#### Lasso

Least absolute shrinkage and selection operation (LASSO) is a well-known method in machine learning, especially for the datasets that have more number of features than number of observations. In regression analysis, LASSO can perform feature selection and regularization at the same time, so as to improve the accuracy of model prediction performance as well as strengthen the interpretability of the obtained model. [21] Lasso could also be applied for classification problem. The function ‘glmnet’ in R (Version: 4.1-1) was used for the LASSO model building, while setting family to ‘binomial’ could build classifiers for the binary outcome.

#### XGBoost

XGBoost is an implementation of the gradient boosted decision tree algorithm. Boosting is also an ensemble algorithm that can combine the output of many weak classifiers into a strong one. The algorithm enables to work on both classification and regression problems. XGBoost, which is defined as a scalable end-to-end tree boosting system, is a very strong boosting method that can be used to build a classifier, and exhibits outstanding prediction performance according to recent studies. [22]. The ‘xgboost’ package in R (Version: 1.4.1.1) allows for applying XGBoost in the classification problem in R.

### Performance Evaluation

The prediction performance of models based on random forest [20], LASSO [21] and XGBoost [22] were evaluated and compared by using 10-fold cross validation. The merged dataset was randomly divided into 10 folds. Each time, the model was built on a combination of the 9 folds, and then evaluated on the leave-out fold. The ROC and AUC were calculated to measure the predictive accuracy of a prediction model [33]. The whole cross validation procedure was replicated 10 times and averaged results were reported.

The feature selection results of these machine learning methods were assessed by calculating the probability of each feature being selected, i.e. the proportion of times that the feature was included in the prediction model based on the 10-fold cross validation. The Spearman’s correlation coefficient was calculated to measure similarity in feature selection between every two methods [34]. The Heatmap and correlation plot were generated to visualize and compare the results.

## Results

### Candidate Features

Due to the large number of gene features in a RNA-seq dataset, we focused on immune checkpoint genes and considered a similar set of candidate features as in [14]. Specifically, we considered 26 immune checkpoint genes that had been reported in the literature and were available in the RNA-seq datasets. As those immune checkpoint genes are often co-stimulatory or co-inhibitory, we considered pairwise relations between those gene expressions to capture their interactions [14, 23–26]. Each pairwise relation feature was an indicator function that compares expression levels of two immune checkpoint genes. A total of 135 pairwise relations features were considered as candidate features in our analysis.

### Data Integration

To enhance statistical power, we merged three published datasets, i.e. Van Allen *et al*. [15], *Hugo et al*. [16], *and Riaz et al*. [17] (Table 1), and obtained a combined dataset with 135 subjects. We first assessed the batch effect for data from different sources. Fig 2A shows the hierarchical clustering of samples from those sources based on the original gene expression data. It is clear that samples from the same source were clustered together, suggesting the presence of batch effect. In contrast, Fig 2B shows the hierarchical clustering of samples based on the 135 features charaterizing the pairwise relations of immune checkpoint genes. Samples from different sources were intermixed. Thus, by focusing on those pairwise relations features, the batch effect was minimized. This is likely due to the fact that pairwise-relation features focus on the relative orders between expressions of pairs of genes. Such information is more robust against batch effect compared to the original gene expression levels.

**Fig 2.**
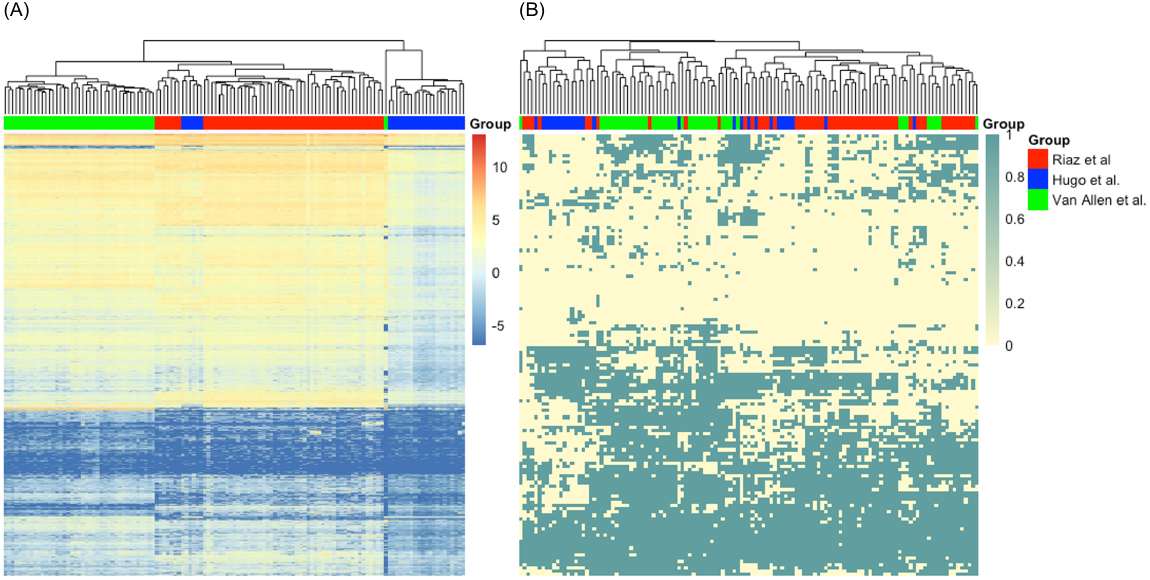
Heatmaps and sample clustering of the merged datasets. based on (A) the original 18878 features and (B) the 135 features charaterizing the pairwise relations of immune checkpoint genes.

### Model Prediction Results

We applied three frequently used statistical/machine learning methods, including random forest [20], least absolute shrinkage and selection operation (LASSO) [21], and XGBoost [22], to build prediction models. Fig 3 presents the ROC curves and AUCs of the prediction models built by the above-mentioned machine learning methods based on 10-fold cross validation. Random forest, and LASSO both had AUCs above 0.7, providing good predictions of the immune response. The XGBoost had a lower AUC of 0.667. The difference in prediction performance across methods is likely due to different feature selection or model building strategies of these methods.

**Fig 3.**
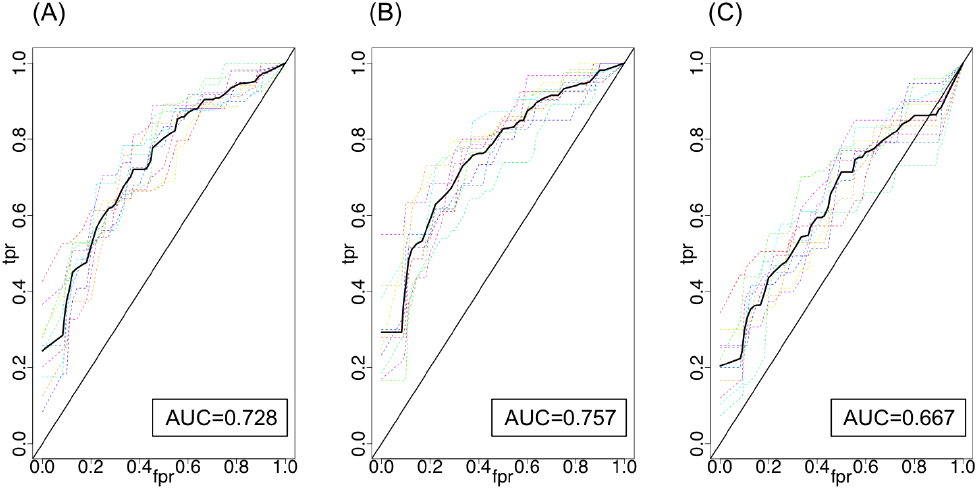
ROC curves of the combined dataset for all algorithms. Comparison of ROC curves among Random Forest (Fig 3A), LASSO (Fig 3B), and XGBoost (Fig 3C) based on the combined dataset. Each colored dashed curve indicates one 10-fold cross validation replicate. The solid black curve indicates the average curve across ten replicates. Results were averaged across ten 10-fold cross-validations. The average area under the curve (AUC) were calculated over the ten replicates.

We next investigated the impact on model’s predictive accuracy by using the combined dataset versus using a single dataset. We applied the same three machine learning methods to build prediction models based on each of the three individual datasets. As shown in Table 2, the AUC from a single dataset was lower than that from the combined dataset. For example, the LASSO AUCs from the single datasets were 0.547, 0.464, and 0.729, respectively, while that from the combined dataset was 0.757. In addition, the ROC curves from different cross-validation replicates had much larger variations (Figure in S1 Figure, Figure in S2 Figure, and Figure in S3 Figure), suggesting that the prediction performance of models based on a single dataset is less stable than that based on the combined dataset. Further, the AUCs from the Hugo et al. dataset tended to be lower than those from the other two datasets for a given machine learning method. This is likely due to the fact that the Hugo et al. dataset had a smaller sample size compared to the other two datasets, which further demonstrates the importance of sample size in prediction model development.

**Table 2.** Summary of perdiction performance.

|  | Build models based on the<br>135 pairwise relations features<br>of immune checkpoint genes |  |  | Build models based on the<br>original 18878 gene<br>expression features |  |  |
| --- | --- | --- | --- | --- | --- | --- |
|  | Random forest | Lasso | XGBoost | Random forest | Lasso | XGBoost |
|  | AUC | AUC | AUC | AUC | AUC | AUC |
| Combined | 0.728 | 0.757 | 0.667 | 0.595* | 0.552* | 0.582* |
| Van Allen et al. | 0.723 | 0.547 | 0.607 | 0.410 | 0.444 | 0.446 |
| Hugo et al. | 0.559 | 0.464 | 0.445 | 0.671 | 0.559 | 0.322 |
| Riaz et al. | 0.711 | 0.729 | 0.622 | 0.568 | 0.441 | 0.447 |

Models were built using Random forest, Lasso, XGBoost based on the combined dataset or an individual dataset. AUCs were calculated based on 10-fold cross validation except for the case of using the Hugo et al. dataset alone, where a 5-fold cross validation was performed because the sample size of the dataset was so small that the 10-fold validation did not yield robust result. \* Data had been normalized based on the Combat method [30,35] when merging the three datasets.

In addition, we assessed the value of incorporating prior biological information and focusing on immune checkpoint genes. As a comparison, we applied the three machine learning methods and feature selection/model building procedure to the original 18878 gene expression features based on either a single dataset or the combined dataset. For the combined dataset, the Combat normalization method had been applied to remove batch effect [30, 35]. Table 2 shows that the AUCs of those resulting models were only around 0.5, even for using the combined dataset. Thus, those models based on original gene expression features without incorporating prior biological knowledge had much poorer prediction performance compared to models using pairwise relationships of immune checkpoint genes. The result indicates that by suggesting biologically relevant features and their combinations, prior biological knowledge can contribute to building better performed prediction models.

### Feature Selection Results

We also compared the features selected by the three statistical/machine learning methods. For each method, the probability of a feature being selected was calculated based on the cross-validation procedure. Features with selection probabilities greater or equal to 0.2 from at least one of the three methods are presented in Fig 4A. Some features, such as ‘PD-1 > PDL-1’, ‘PD-1 > CTLA4’, ‘PD-1 > CD200R1’, ‘PD-1 > TNFRSF18’, ‘PD-1 > CD137L’, ‘PDL-1 > CTLA4’, ‘CTLA4 > CD200R1’, ‘CD80 > CD137L’, ‘CD86 > IL2RB’, tended to be selected by all the methods with similar probabilities, while some other features had very different selection probabilities for different methods. A table with detailed records of probabilities is provided in Table in S1 Table.

**Fig 4.**
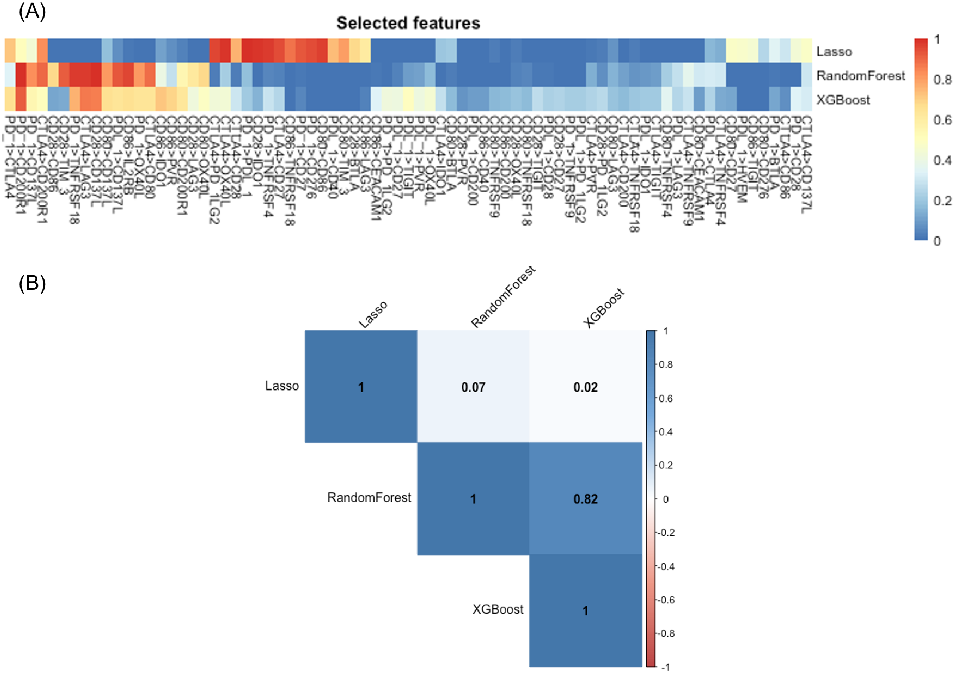
Feature selection results. (A) Comparison of feature selection probabilities across LASSO, random forest and XGBoost. Only features with probability of selection larger or equal to 0.2 from at least one of the three methods are presented. (B) Spearman correlation of feature selection probabilities between each pair of methods.

We further quantified the consistency in feature selection between each pair of methods based on the Spearman correlation coefficient. Results are presented in Fig 4B. The two tree-based methods, random forest and XGBoost, had high consistency in feature selection. The Spearman’s correlation coefficients was around 0.82. In contrast, the features selected by LASSO were very different from the tree-based methods with the Spearman’s correlation coefficients less than 0.1. This is likely due to the fact that tree-based methods focus on non-linear combinations while LASSO focuses on linear combinations across features.

## Discussion

We considered three frequently used statistical/machine learning methods to construct prediction models. For all three methods, models based on merged data had higher predictive accuracy than those based on individual datasets. This result suggested that the improved prediction performance is not sensitive to the choice of model construction method. We also noticed some difference in predictive accuracy between models from different machine learning methods. Further investigation of methods’ predictive accuracy under different sample size settings will be needed to more comprehensively evaluate and compare the performance of different methods for predicting ICB treatment response in melanoma patients.

We focused on interactions between immune checkpoint genes as candidate features and followed Auslander et. al to use logical relations between the expression levels of pairs of immune checkpoint genes as candidate features to characterize interactions between those genes [14]. One can consider other function forms, e.g. products of expression levels of pairs of genes, to describe the co-stimulatory and co-inhibitory effects. An interesting topic for future research is to compare different function forms and identify more informative function forms to enhance prediction.

Removing batch effect is an important task when merging different datasets. We showed that features on pairwise relations between immune checkpoint genes were less affected by batches compared to the original features. This is because the pairwise relation features only consider the order of expressions between genes but not the quantitative expression levels. Therefore, focusing on pairwise relation features reduces the impact of batch effect in our analysis. We also tried a more traditional approach, ComBat [30, 35], for removing batch effect. However, we noticed that the ComBat adjusted expression values did not pertain the order of expressions between genes. Therefore, such batch effect removal may cause disturbance of the useful information contained in the original data.

The ICB therapy for melanoma primarily targets cytotoxic T-lymphocyte-associated protein 4 (CTLA-4) and programmed cell death protein 1 (PD-1) [27]. Both CTLA-4 and PD-1 binding have similar negative regulatory effects on the activity of T-cells. Anti-PD-1 and Anti-CTLA-4 immunotherapies inhibit these targets and prevent melanoma cells from evading the immune system [27]. In the literature, it has been shown that gene expression signatures can be generally applicable to predict treatment effects of both anti-CTLA-4 and anti-PD-1 drugs [13, 14]. Therefore, in this paper, we combined datasets for drugs targeting CTLA-4 and datasets for drugs targeting PD-1 to ensure an adequate sample size for model building [15–17]. In the future, as clinical trial data acumulate, one can possibly focus on trials for drugs targeting one of the two targets to develop target-specific gene expression signatures. It would be interesting to investigate whether those gene signatures could further improve the predictive accuracy of treatment effect.

## Conclusion

In summary, we have demonstrated that merging datasets and incorporating prior biological knowledge are useful strategies to improve the prediction performance of ICB treatment using gene expression signatures. The batch effect could be minimized by capturing pairwise-relation features. Classical machine learning algorithms were applied to the integrated datasets with features of pairwise relations, and demonstrated satisfactory classification performance, with AUC around 0.70. When compared with the model built on the single dataset, the result showed that the model with dataset merging improved and stabilized the prediction performance. In addition, the prediction performance of models based on the pairwise relations of immune checkpoint genes was higher than models built on the original dataset without incorporating prior biological knowledge. Overall, our finding demonstrated that merging datasets from multiple studies and incorporating prior biological knowledge are of high value to identify ICB response biomarkers in future studies.

## Data Availability

All data produced in the present study are available upon reasonable request to the authors.

## Ethical Approval and Consent to participate

This study is approved by the University of Kentucky Institutional Review Board.

## Human Ethics

Not applicable

## Consent for publication

Not applicable

## Availability of supporting data

The supporting data can be available to public.

## Acknowledgments

The authors acknowledge Daheng He for the help in RNA-seq data processing.

## Supporting information

### List of feature selection and probability

**Table 3.** Feature selection with different algorithms

| Index | Variable Name | Random Forest | Lasso | XGBoost |
| --- | --- | --- | --- | --- |
| 1 | PD-1>PDL-1 | 0.18 | 1 | 0.21 |
| 2 | PD-1>CTLA4 | 0.34 | 0.72 | 0.66 |
| 3 | PD-1>CD28 | 0 | 0.44 | 0.34 |
| 4 | PD-1>CD80 | 0.01 | 0 | 0.14 |
| 5 | PD-1>CD86 | 0 | 0 | 0.04 |
| 6 | PD-1>BTLA | 0.02 | 0.35 | 0.12 |
| 7 | PD-1>CD200 | 0 | 0 | 0.01 |
| 8 | PD-1>CD200R1 | 1 | 0.51 | 0.92 |
| 9 | PD-1>CD27 | 0.02 | 0.93 | 0.05 |
| 10 | PD-1>CD276 | 0 | 0.95 | 0 |
| 11 | PD-1>CD40 | 0 | 0 | 0 |
| 12 | PD-1>CEACAM1 | 0 | 0.03 | 0.02 |
| 13 | PD-1>TIM-3 | 0 | 0.02 | 0 |
| 14 | PD-1>IDO1 | 0 | 0.01 | 0.02 |
| 15 | PD-1>IL2RB | 0 | 0.02 | 0 |
| 16 | PD-1>LAG3 | 0 | 0 | 0.03 |
| 17 | PD-1>PD-1LG2 | 0 | 0 | 0.42 |
| 18 | PD-1>PVR | 0 | 0.07 | 0.05 |
| 19 | PD-1>TIGIT | 0 | 0.05 | 0.04 |
| 20 | PD-1>HVEM | 0 | 0.01 | 0.06 |
| 21 | PD-1>TNFRSF18 | 0.98 | 0.01 | 0.76 |
| 22 | PD-1>TNFRSF4 | 0.05 | 0.98 | 0.29 |
| 23 | PD-1>TNFRSF9 | 0 | 0.01 | 0.22 |
| 24 | PD-1>OX40L | 0.81 | 0 | 0.64 |
| 25 | PD-1>CD137L | 0.83 | 0.44 | 0.55 |
| 26 | PDL-1>CTLA4 | 0.32 | 0.16 | 0.24 |
| 27 | PDL-1>CD28 | 0.02 | 0 | 0.23 |
| 28 | PDL-1>CD80 | 0 | 0.06 | 0 |
| 29 | PDL-1>CD86 | 0.01 | 0 | 0.19 |
| 30 | PDL-1>BTLA | 0 | 0 | 0 |
| 31 | PDL-1>CD200 | 0.01 | 0 | 0.1 |
| 32 | PDL-1>CD200R1 | 0 | 0.01 | 0.02 |
| 33 | PDL-1>CD27 | 0.04 | 0.01 | 0.41 |
| 34 | PDL-1>CD276 | 0 | 0.04 | 0 |
| 35 | PDL-1>CD40 | 0 | 0.74 | 0 |
| 36 | PDL-1>CEACAM1 | 0 | 0.13 | 0.02 |
| 37 | PDL-1>TIM-3 | 0 | 0 | 0 |
| 38 | PDL-1>IDO1 | 0.09 | 0 | 0.21 |
| 39 | PDL-1>IL2RB | 0 | 0 | 0.07 |
| 40 | PDL-1>LAG3 | 0.28 | 0 | 0.25 |
| 41 | PDL-1>PD-1LG2 | 0 | 0 | 0.22 |
| 42 | PDL-1>PVR | 0.11 | 0 | 0.41 |
| 43 | PDL-1>TIGIT | 0.1 | 0 | 0.5 |
| 44 | PDL-1>HVEM | 0 | 0.47 | 0 |
| 45 | PDL-1>TNFRSF18 | 0 | 0.01 | 0.03 |
| 46 | PDL-1>TNFRSF4 | 0.11 | 0 | 0.1 |
| 47 | PDL-1>TNFRSF9 | 0 | 0 | 0 |
| 48 | PDL-1>OX40L | 0.17 | 0 | 0.45 |
| 49 | PDL-1>CD137L | 0.93 | 0.07 | 0.64 |

Table 3 – continued from previous page
| Index | Variable Name | Random Forest | Lasso | XGBoost |
| --- | --- | --- | --- | --- |
| 50 | CTLA4>CD28 | 0.03 | 0.76 | 0.3 |
| 51 | CTLA4>CD80 | 0.89 | 0 | 0.64 |
| 52 | CTLA4>CD86 | 0 | 0.3 | 0.07 |
| 53 | CTLA4>BTLA | 0 | 0 | 0.04 |
| 54 | CTLA4>CD200 | 0.11 | 0 | 0.17 |
| 55 | CTLA4>CD200R1 | 0.9 | 0.83 | 0.51 |
| 56 | CTLA4>CD27 | 0.04 | 0.95 | 0.25 |
| 57 | CTLA4>CD276 | 0 | 0 | 0 |
| 58 | CTLA4>CD40 | 0 | 0 | 0.02 |
| 59 | CTLA4>CEACAM1 | 0 | 0 | 0.02 |
| 60 | CTLA4>TIM-3 | 0 | 0 | 0.04 |
| 61 | CTLA4>IDO1 | 0.01 | 0.2 | 0.28 |
| 62 | CTLA4>IL2RB | 0 | 0.01 | 0.09 |
| 63 | CTLA4>LAG3 | 0.98 | 0 | 0.87 |
| 64 | CTLA4>PD-1LG2 | 0.03 | 0.96 | 0.43 |
| 65 | CTLA4>PVR | 0.14 | 0.02 | 0.24 |
| 66 | CTLA4>TIGIT | 0.1 | 0 | 0.21 |
| 67 | CTLA4>HVEM | 0 | 0.05 | 0 |
| 68 | CTLA4>TNFRSF18 | 0.08 | 0.03 | 0.2 |
| 69 | CTLA4>TNFRSF4 | 0.31 | 0.14 | 0.13 |
| 70 | CTLA4>TNFRSF9 | 0.37 | 0.01 | 0.29 |
| 71 | CTLA4>OX40L | 0.22 | 0.99 | 0.44 |
| 72 | CTLA4>CD137L | 0.3 | 0.48 | 0.32 |
| 73 | CD28>CD80 | 0 | 0 | 0.13 |
| 74 | CD28>CD86 | 0.68 | 0.01 | 0.12 |
| 75 | CD28>BTLA | 0 | 0.68 | 0.01 |
| 76 | CD28>CD200 | 0 | 0.08 | 0.03 |
| 77 | CD28>CD200R1 | 0 | 0 | 0.06 |
| 78 | CD28>CD27 | 0 | 0.01 | 0.24 |
| 79 | CD28>CD276 | 0 | 0.04 | 0 |
| 80 | CD28>CD40 | 0 | 0 | 0 |
| 81 | CD28>CEACAM1 | 0 | 0.1 | 0.02 |
| 82 | CD28>TIM-3 | 0.92 | 0.01 | 0.13 |
| 83 | CD28>IDO1 | 0.01 | 0.99 | 0.15 |
| 84 | CD28>IL2RB | 0 | 0 | 0.09 |
| 85 | CD28>LAG3 | 0.62 | 0.01 | 0.42 |
| 86 | CD28>PD-1LG2 | 0.07 | 0.03 | 0.26 |
| 87 | CD28>PVR | 0 | 0.01 | 0.08 |
| 88 | CD28>TIGIT | 0.02 | 0 | 0.27 |
| 89 | CD28>HVEM | 0 | 0 | 0 |
| 90 | CD28>TNFRSF18 | 0 | 0 | 0.02 |
| 91 | CD28>TNFRSF4 | 0.01 | 0 | 0.2 |
| 92 | CD28>TNFRSF9 | 0 | 0.03 | 0.07 |
| 93 | CD28>OX40L | 0.04 | 0.03 | 0.19 |
| 94 | CD28>CD137L | 1 | 0 | 0.86 |
| 95 | CD80>CD86 | 0 | 0.97 | 0 |
| 96 | CD80>BTLA | 0.03 | 0.21 | 0.17 |
| 97 | CD80>CD200 | 0 | 0.01 | 0 |
| 98 | CD80>CD200R1 | 0.55 | 0.04 | 0.63 |
| 99 | CD80>CD27 | 0.01 | 0.48 | 0.12 |
| 100 | CD80>CD276 | 0 | 0.25 | 0 |
| 101 | CD80>CD40 | 0 | 0 | 0 |
| 102 | CD80>CEACAM1 | 0.31 | 0 | 0.1 |
| 103 | CD80>TIM-3 | 0 | 0.8 | 0 |

**Table 3 – continued from previous page**
| **Index** | **Variable Name** | **Random Forest** | **Lasso** | **XGBoost** |
| --- | --- | --- | --- | --- |
| 104 | CD80>IDO1 | 0.03 | 0 | 0.11 |
| 105 | CD80>IL2RB | 0 | 0.04 | 0 |
| 106 | CD80>LAG3 | 0.15 | 0.08 | 0.19 |
| 107 | CD80>PD-1LG2 | 0.04 | 0 | 0.17 |
| 108 | CD80>PVR | 0 | 0.05 | 0.02 |
| 109 | CD80>TIGIT | 0 | 0.06 | 0.11 |
| 110 | CD80>HVEM | 0 | 0 | 0.08 |
| 111 | CD80>TNFRSF18 | 0.05 | 0 | 0.17 |
| 112 | CD80>TNFRSF4 | 0.22 | 0 | 0.36 |
| 113 | CD80>TNFRSF9 | 0.01 | 0.03 | 0.16 |
| 114 | CD80>OX40L | 0.68 | 0.01 | 0.49 |
| 115 | CD80>CD137L | 0.81 | 0.18 | 0.66 |
| 116 | CD86>BTLA | 0 | 0.09 | 0 |
| 117 | CD86>CD200 | 0 | 0.01 | 0.17 |
| 118 | CD86>CD200R1 | 0 | 0.01 | 0 |
| 119 | CD86>CD27 | 0 | 0 | 0.07 |
| 120 | CD86>CD276 | 0 | 0.01 | 0 |
| 121 | CD86>CD40 | 0.01 | 0 | 0.1 |
| 122 | CD86>CEACAM1 | 0.02 | 0 | 0.34 |
| 123 | CD86>TIM-3 | 0.01 | 0 | 0.06 |
| 124 | CD86>IDO1 | 0.4 | 0.11 | 0.72 |
| 125 | CD86>IL2RB | 0.96 | 0.02 | 0.66 |
| 126 | CD86>LAG3 | 0 | 0.61 | 0.04 |
| 127 | CD86>PD-1LG2 | 0 | 0.01 | 0 |
| 128 | CD86>PVR | 0.28 | 0.07 | 0.69 |
| 129 | CD86>TIGIT | 0 | 0.45 | 0 |
| 130 | CD86>HVEM | 0 | 0.02 | 0 |
| 131 | CD86>TNFRSF18 | 0 | 0.88 | 0 |
| 132 | CD86>TNFRSF4 | 0 | 0 | 0.01 |
| 133 | CD86>TNFRSF9 | 0 | 0.04 | 0 |
| 134 | CD86>OX40L | 0 | 0 | 0 |
| 135 | CD86>CD137L | 0.01 | 0.01 | 0.03 |

### ROC curves

**Fig 5.**
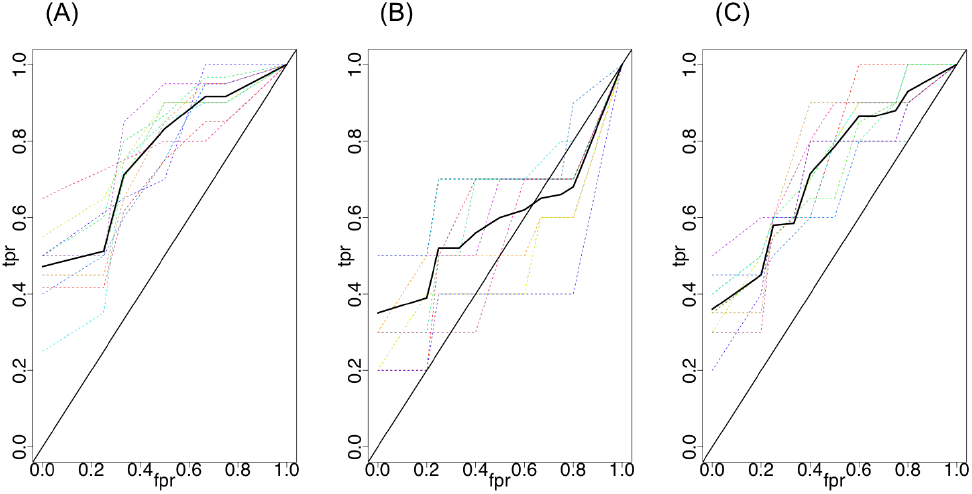
Random forest - ROC plot for the comparison of the result. Comparison of ROC curves with applying Random Forest on each single dataset. (A) Cross validation using single dataset Van Allen et al.; (B) Cross validation using single dataset Hugo et al.; (C) Cross validation using single dataset Riaz et al. Each colored dashed curve indicates one 10-fold cross validation replicate. The solid black curve indicates the average curve across ten replicates. Results were averaged across ten 10-fold cross-validations. The average area under the curve (AUC) were calculated over the ten replicates.

**Fig 6.**
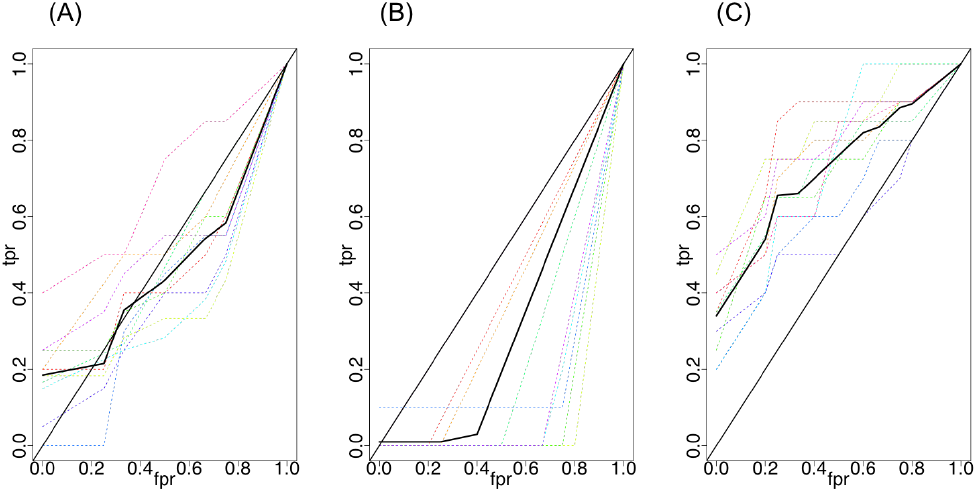
Lasso - ROC plot for the comparison of the result. Comparison of ROC curves with applying Lasso on each single dataset. (A) Cross validation using single dataset Van Allen et al.; (B) Cross validation using single dataset Hugo et al.; (C) Cross validation using single dataset Riaz et al.

Some of the Lasso ROC curves were not present, since some of the models did not select any RNA-seq pairs, with only intercept left and leading to an AUC of 0.5.

Similarly, using XGBoost algorithm, some of the models did not select any feature, and thus non-tree model was detected. Hence, some ROC curves were not shown in Fig 7 (middle panel).

**Fig 7.**
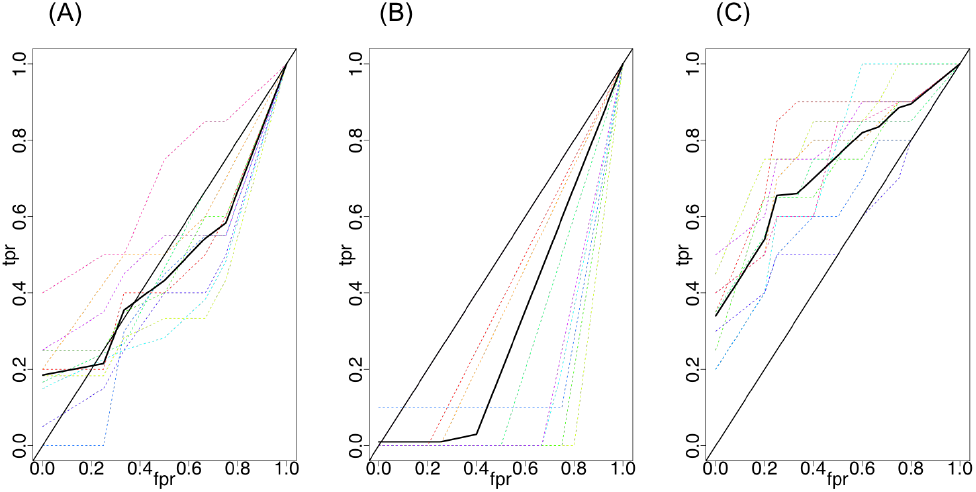
XGBoost - ROC plot for the comparison of the result. Comparison of ROC curves with applying XGBoost on each single dataset. (A) Cross validation using single dataset Van Allen et al.; (B) Cross validation using single dataset Hugo et al.; (C) Cross validation using single dataset Riaz et al.

